# Validation of an open-source smartphone step counting algorithm in clinical and non-clinical settings

**DOI:** 10.1101/2023.03.28.23287844

**Authors:** Marcin Straczkiewicz, Nancy L. Keating, Embree Thompson, Ursula A. Matulonis, Susana M. Campos, Alexi A. Wright, Jukka-Pekka Onnela

**Author notes:** Corresponding author: Marcin Straczkiewicz. Denotes shared last authorship (Alexi A. Wright, Jukka-Pekka Onnela).

## Abstract

**Background:** Step counts are increasingly used in public health and clinical research to assess wellbeing, lifestyle, and health status. However, estimating step counts using commercial activity trackers has several limitations, including a lack of reproducibility, generalizability, and scalability. Smartphones are a potentially promising alternative, but their step-counting algorithms require robust validation that accounts for temporal sensor body location, individual gait characteristics, and heterogeneous health states.

**Objective:** Our goal was to evaluate an open-source step-counting method for smartphones under various measurement conditions against step counts estimated from data collected simultaneously from different body locations (“internal” validation), manually ascertained ground truth (“manual” validation), and step counts from a commercial activity tracker (Fitbit Charge 2) in patients with advanced cancer (“wearable” validation).

**Methods:** We used eight independent datasets collected in controlled, semi-controlled, and free-living environments with different devices (primarily Android smartphones and wearable accelerometers) carried at typical body locations. Five datasets (N=103) were used for internal validation, two datasets (N=107) for manual validation, and one dataset (N=45) used for wearable validation. In each scenario, step counts were estimated using a previously published step-counting method for smartphones that uses raw sub-second level accelerometer data. We calculated mean bias and limits of agreement (LoA) between step count estimates and validation criteria using Bland-Altman analysis.

**Results:** In the internal validation datasets, participants performed 751.7±581.2 (mean±SD) steps, and the mean bias was -7.2 steps (LoA -47.6, 33.3) or -0.5%. In the manual validation datasets, the ground truth step count was 367.4±359.4 steps while the mean bias was -0.4 steps (LoA -75.2, 74.3) or 0.1 %. In the wearable validation dataset, Fitbit devices indicated mean step counts of 1931.2±2338.4, while the calculated bias was equal to -67.1 steps (LoA -603.8, 469.7) or a difference of 0.3 %.

**Conclusions:** This study demonstrates that our open-source step counting method for smartphone data provides reliable step counts across sensor locations, measurement scenarios, and populations, including healthy adults and patients with cancer.

## Introduction

Walking is the most common form of physical activity (1). It is also important to prevent chronic disease and premature mortality (2–4). The recent proliferation and integration of wearable activity trackers into public health and clinical research studies has allowed investigators to identify gait-related biomarkers, such as decreased daily step counts, as risk factors for cardiovascular disease, cancer, stroke, and type 2 diabetes (5–10).

Despite the potential for wearable activity trackers to increase physical activity, improve health, and provide unique behavioral insights, there are several important limitations. First, the adoption of wearables is uneven across the population, and most people stop using wearable activity trackers after 6 months (11–14). Second, commercial devices rarely allow access to their raw (sub-second level) data, or provide open-source algorithms for processing data into clinically meaningful endpoints (15–17). Third, the accuracy of step count estimates are affected by metrological and behavioral factors, such as the location of the wearable on the body and temporal gait speed (18–20).

Smartphones are a promising alternative for collecting objective, scalable, and reproducible data about human behavior (21–24). Although smartphones can overcome many limitations of wearable activity trackers—e.g., through access to raw sensor data (25) and increased adoption among older individuals (26)— the quantification of gait-related biomarkers remains challenging. This is largely because of the variation in the location and orientation of smartphones in relation to the body in real-life conditions, which affects the data collected from smartphones’ inertial sensors (27–29).

To address this problem, we have recently proposed an open-source walking recognition method (29), which can be applied to accelerometer data collected from various locations on the body, making it suitable for smartphones. In this paper, we demonstrate how our method can be used for quantifying steps, and we validate its performance in eight independent datasets. We validate this method in three ways: (i) by comparing step counts estimated from multiple sensors worn simultaneously at various known body locations; (ii) by comparing step counts estimated from a sensor worn at an unspecified body location against visually assessed and manually annotated ground truth (i.e., observed step counts); and (iii) by comparing step counts estimated from a sensor worn at an unspecified body location against estimates provided by an independent commercial activity tracker (Fitbit Charge 2) worn on the wrist. The first (“internal”) and second (“manual”) validations involve healthy subjects whose data were obtained from publicly available datasets collected in controlled, semi-controlled, and free-living conditions, while the third (“wearable”) validation includes data collected by our team from patients with advanced cancer receiving chemotherapy as outpatients in free-living conditions.

## Methods

### Step counting algorithm

Our method leveraged the observation that regardless of the sensor location, orientation, or subject, during walking activity device’s accelerometer signal oscillates around a local mean with a frequency equal to the performed steps (29). To extract this information, we used the continuous wavelet transform to project the original signal onto the time-frequency space of wavelet coefficients which are maximized when a particular frequency matches the frequency of the observed signal at a given time point (**Figure 1**). To translate this information into number of steps, we split the projection into non-overlapping one-second windows, and we estimated the temporal step frequency as a frequency with the maximum average wavelet coefficient. The estimated frequency reflects the number of steps a person performs within this time window. Finally, the total number of steps was calculated as a sum of all one-second step counts calculated over the duration of the observed period of walking.

**Figure 1.**
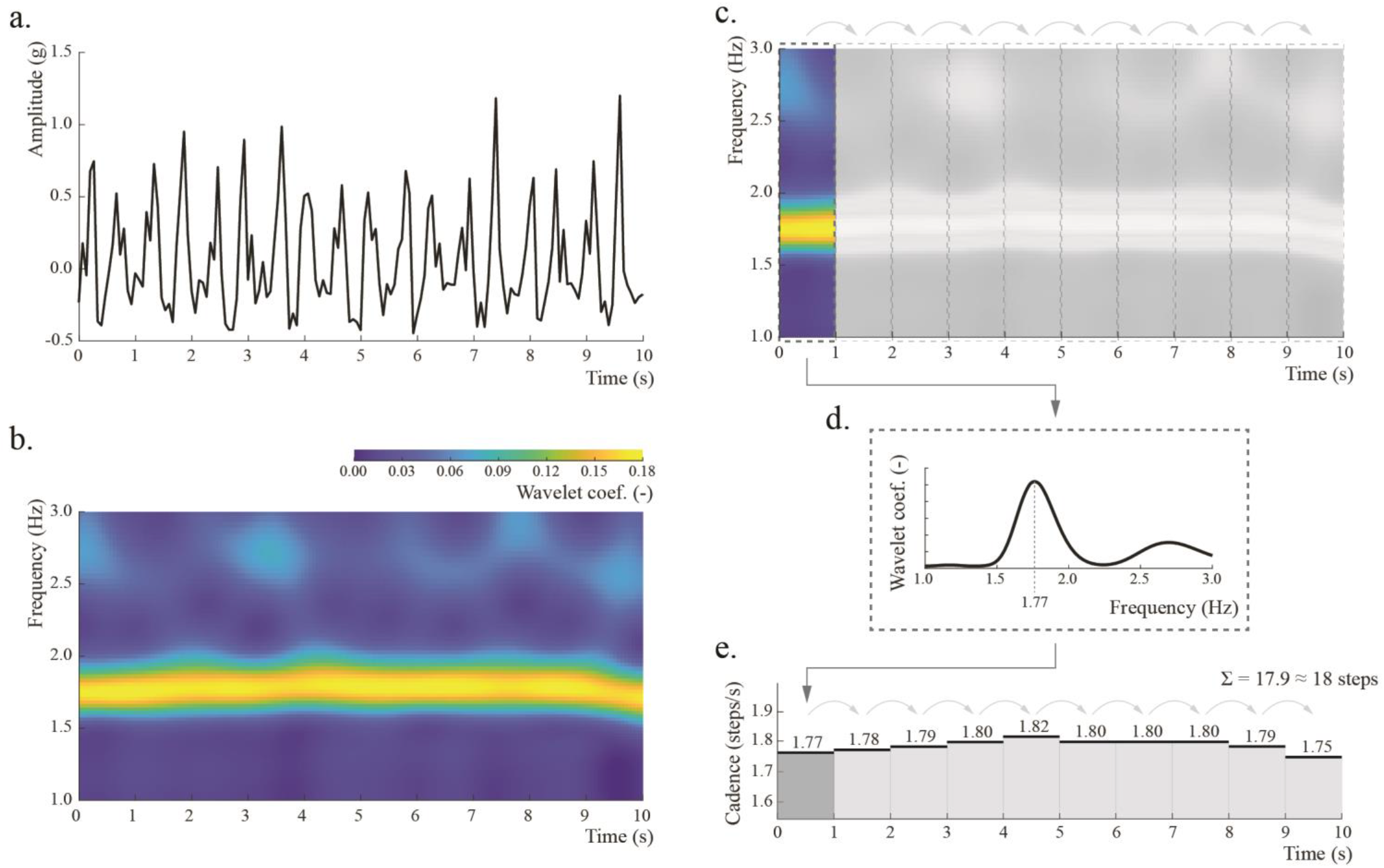
The step counting algorithm. The original signal (a.) is projected onto the time-frequency space (b.) using wavelet transformation, which shows the relative weights of different frequencies over time (brighter color indicates higher weight). This scalogram is then split into non-overlapping one-second windows (c.). The temporal step frequency (cadence) is estimated as a frequency with maximum average wavelet coefficient inside each window (d.). The total number of steps in a signal is calculated as a rounded sum of all one-second counts in that signal (e.).

The step counting method described above is embedded into the walking recognition algorithm published in the public domain (30,31).

### Data description

We evaluated the step counting method in three ways, which we refer to as “internal,” “manual,” and “wearable” validations. Each approach was selected to assess a different aspect of the method’s performance: the internal validation aimed to determine the consistency of step counts across different body locations; the manual validation aimed to assess the method’s accuracy against step counts assessed visually by an observer; and the wearable validation aimed to assess the method’s step count compared with step counts obtained from a commercial, consumer-grade activity tracker (Fitbit Charge 2) worn at the wrist. Cumulatively, the entire validation was conducted using eight independent datasets, including seven datasets available in the public domain and one dataset collected by our research team. All datasets are described in the following subsections.

#### Internal validation

For the internal validation, we identified five publicly available datasets, including: Daily Life Activities (*DaLiAc*) (32), Physical Activity Recognition Using Smartphone Sensors (*PARUSS*) (33), Real-World (*RealWorld*) (34), Simulated Falls and Daily Living Activities (*SFDLA*) (35), and Human Physical Activity (*SPADES*) (36). The datasets contained accelerometer data on walking activity collected simultaneously at several body locations that are representative of everyday use of smartphones.

The aggregated internal validation dataset included measurements collected from 103 healthy adults (**Table 1**) who performed walking activities in controlled environments (i.e., all participants followed some predefined path), typically around a university campus (**Table 2**). One dataset, *RealWorld*, involved participants walking outside in a parking lot and a forest.

**Table 1.** Demographics, body measures, and health status of participants involved in datasets included in this study. Age, height, weight, and BMI are provided as range (mean±SD), when available.

| Validation scheme | Dataset | N | Sex | Age | Height | Weight | BMI | Health status |
| --- | --- | --- | --- | --- | --- | --- | --- | --- |
|  |  |  | N<br>male | years | cm | kg | kg/m<sup>2</sup> |  |
| **Internal** |  |  |  |  |  |  |  |  |
|  | **DaLiAc** |  |  |  |  |  |  |  |
| | | 19 | 11 | 18-55<br>(26.5±7.7) | 158-196<br>(177.0±11.1) | 54-108<br>(75.2±14.2) | 17-34<br>(23.9±3.7) | Healthy |
|  | **PARUSS** |  |  |  |  |  |  |  |
|  |  | 10 | 10 | 25-30 | N/A | N/A | N/A | Healthy |
|  | **RealWorld** |  |  |  |  |  |  |  |
| | | 15 | 8 | 16-62<br>(31.9±12.4) | 163-183<br>(173.1±6.9) | 48-95<br>(74.1±13.8) | 18-35<br>(24.7±4.4) | Healthy |
|  | **SFDLA** |  |  |  |  |  |  |  |
| | | 17 | 10 | 19-27<br>(21.9±2.0) | 157-184<br>(171.6±7.8) | 47-92<br>(65.0±13.9) | 17-31<br>(21.9±3.7) | Healthy |
|  | **SPADES** |  |  |  |  |  |  |  |
| | | 42 | 27 | 18-30<br>(23.5±3.1) | 151-180<br>(174.2±8.5) | 51-112<br>(73.8±15.0) | 18-35<br>(24.7±4.1) | Healthy |
| **Manual** |  |  |  |  |  |  |  |  |
|  | **WalkRec** |  |  |  |  |  |  |  |
|  |  | 77 | N/A | N/A | N/A | N/A | N/A | Healthy |
|  | **PedEval** |  |  |  |  |  |  |  |
| | | 30 | 15 | 19-27<br>(21.9±52.4) | 152-193<br>(171.0±10.8) | 43-136<br>(70.5±17.6) | 17-37<br>(23.8±3.7) | Healthy |
| **Wearable** |  |  |  |  |  |  |  |  |
|  | HOPE |  |  |  |  |  |  |  |
|  |  | 45 | 0 | 24-79<br>(61.5±11.8) | 148-172<br>(159.9±6.1) | 48-107<br>(67.8±13.0) | 19-43<br>(26.5±4.9) | Patients<br>with<br>advanced<br>cancer |

**Table 2.**
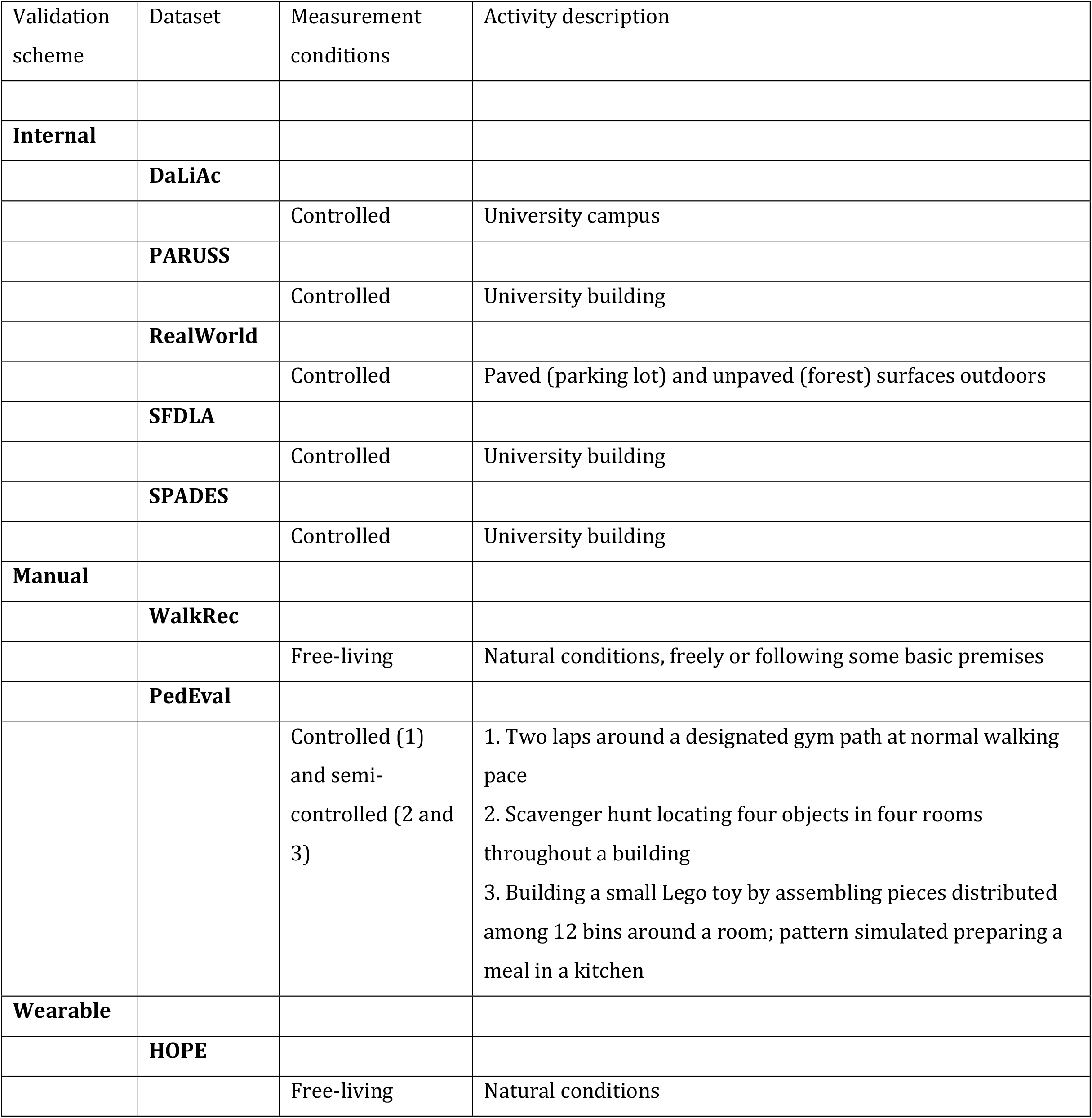
Walking conditions in the datasets included in this study.

Accelerometer data were collected using various wearable devices, including Android-based smartphones and research-grade wearable accelerometers from SHIMMER (Dublin, Ireland), Xsens Technologies (Enschede, The Netherlands), and ActiGraph (Pensacola, FL). The devices were positioned at various locations across the body, i.e., around the thigh, at the waist, on the chest, and on the arm (**Table 3**). Dataset measurements differed based upon data collection parameters, including the sampling frequency (e.g., between 25 Hz in *SFDLA* to 204.8 Hz in *DaLiAc*) and measurement range (between ±6 g in *DaLiAc* to ±12 g in *SFDLA*). The measurement range was not provided in the *PARUSS* and *RealWorld* datasets.

**Table 3.** Measurement parameters for the datasets included in this study.

| Validation scheme | Dataset | Sensing device | Sensor location | Measurement range | Sampling frequency |
| --- | --- | --- | --- | --- | --- |
|  |  |  |  | g | Hz |
| **Internal** |  |  |  |  |  |
|  | **DaLiAc** |  |  |  |  |
| | | Wearable accelerometer: SHIMMER | Waist, chest | $\pm 6$ | 204.8 |
|  | **PARUSS** |  |  |  |  |
|  |  | Smartphone: Samsung Galaxy S2 | Thigh, waist, arm | N/A | 50 |
|  | **RealWorld** |  |  |  |  |
|  |  | Smartphone: Samsung Galaxy S4 | Thigh, waist, chest, arm | N/A | 50 |
|  | **SFDLA** |  |  |  |  |
| | | Wearable accelerometer: Xsens MTw | Thigh, waist, chest | $\pm 12$ | 25 |
|  | **SPADES** |  |  |  |  |
| | | Wearable accelerometer: ActiGraph GT9X | Thigh, waist | $\pm 8$ | 80 |
| **Manual** |  |  |  |  |  |
|  | **WalkRec** |  |  |  |  |
|  |  | Smartphone: BQ Aquaris M5 | Unspecified | N/A | 100 |
|  | **PedEval** |  |  |  |  |
| | | Wearable accelerometer: SHIMMER3 | Waist | $\pm 4$ | 15 |
| **Wearable** |  |  |  |  |  |
|  | **HOPE** |  |  |  |  |
|  |  | Smartphone: various Android- and iOS-based | Unspecified | Various | Various |

#### Manual validation

Manual validation was performed using two publicly available datasets: Walking Recognition (*WalkRec*) (37) and the Pedometer Evaluation Project (*PedEval*) (38). The aggregated dataset consisted of both raw accelerometer data for 107 healthy participants and ground truth step counts for each walking activity performed by study participants.

In this approach, walking activities were performed in controlled, semi-controlled, or free-living conditions. Specifically, *WalkRec* participants walked in settings of their choice without specific instructions; e.g. indoor and outdoor walking along flat surfaces, climbing stairs, etc. (free-living), while *PedEval* participants performed three prescribed walking tasks: (1) a two-lap stroll along a designated path (controlled), (2) a scavenger hunt across four rooms (semi-controlled), and (3) a toy-assembling assignment using pieces distributed across a dozen bins located around a room (semi-controlled). In *PedEval*, step counts were visually assessed and manually annotated by a research team member, while in *WalkRec*, the ground truth annotation was further augmented by recordings from a separate smartphone placed on each participant’s ankle.

The manual validation dataset was collected either by Android-based smartphones or a wearable accelerometer (SHIMMER3) placed around the waist (*PedEval*) or at various unspecified locations across the body (*WalkRec)*. Each dataset was collected with a different sampling frequency (*WalkRec* 15 Hz, *PedEval* 100 Hz), and only *PedEval* provided a measurement range (±4 g).

#### Wearable validation

The wearable validation dataset was collected from patients with advanced gynecologic cancers receiving outpatient chemotherapy as part of the Helping Our Patients Excel (*HOPE)* study. The *HOPE* study aimed to assess the feasibility, acceptability, and perceived effectiveness of a mobile health intervention that used commercial wearable activity trackers and Beiwe, a digital phenotyping research platform, to collect accelerometer data, smartphone sensor data and patient-reported outcomes. Patients were recruited from the outpatient gynecological oncology clinic at the Dana-Farber Cancer Institute in Boston, MA. The inclusion and exclusion criteria for study participation are described elsewhere (39).

The dataset included 45 female patients with recurrent gynecologic cancers, including ovarian (n=34), uterine (n=5), cervical (n=5) and vulvar (n=1) cancers. Patients were asked to wear the Fitbit Charge 2 (Fitbit, San Francisco, CA) on their nondominant wrist during all waking hours for a period of six months in a free-living setting. Each Fitbit was linked to the Fitabase analytics system (Small Steps Laboratories, San Diego, CA), which enabled the investigators to remotely monitor and export several metrics of patients’ physical activity, including minute-level step counts.

At baseline, patients were also asked to install Beiwe, the front-end component of the open-source, high-throughput digital phenotyping platform designed and maintained by members of the Harvard T.H. Chan School of Public Health (40). Among other passive data streams, Beiwe collected raw accelerometer data with the default sampling rate (typically 10 Hz on most phones) using a sampling scheme that alternated between on-cycles and off-cycles, corresponding to time intervals when the sensor actively collected data and was dormant, respectively. The smartphone’s accelerometer was configured to follow a 10-second on-cycle and 20-second off-cycle. The sample scheme was identical on all participants’ smartphones.

### Data preprocessing

Because each dataset had different data collection parameters, we preprocessed datasets to standardize the inputs in our algorithm. First, we verified if the acceleration data were provided in gravitational units (g); data provided in SI units were converted using the standard definition 1 g = 9.80665 m/s^2^. Second, we used linear interpolation to impose a uniform sampling frequency of 10 Hz across tri-axial accelerometer data. Third, we transformed the tri-axial accelerometer signals into sensor orientation-invariant vector magnitude.

### Statistical Analysis

The available accelerometer data were processed using the walking recognition and step counting algorithm with default tuning parameters, as previously described (29). Depending on the validation approach, the resulting one-second step counts were then aggregated into step counts for the entire walking bout or specified time fragment. For the internal and manual validations, step counts were calculated as a sum of all step counts in each walking bout and for each sensor location.

Additional analyses were required for the wearable validation. Here, steps counts were first aggregated on a minute level, the smallest time resolution available to export from Fitabase. Because the Beiwe sampling scheme follows on- and off-cycles, we adjusted the observed smartphone-based step counts by a proportional recovery based on the ratio between the duration of data collection (20 s) and non-collection (40 s) in each one-minute window by multiplying them by 3. Further, due to lack of information on both wearable and smartphone wear-time, but also a potential time-lag between measurements between the two devices, we removed minutes with 0 steps recorded by either method. Finally, to allow for a direct comparison, we summed the smartphone-based step counts for each day of observation.

Each validation procedure considered a different ground truth step count for comparison. In the internal validation sample, we compared step counts estimated from various body locations for the entire walking bout. For example, if the dataset included data from three sensors located on the thigh, waist, and arm, we compared step counts between the thigh and waist, thigh and arm, and waist and arm. In the manual validation sample, we compared step counts estimated from the available sensor location to a visually assessed ground truth for the entire walking bout. In the wearable validation sample, we compared daily number of steps estimated from the smartphone to step counts provided by Fitbit. This procedure was performed using two days of observations for each patient. The first day was identified as the first full day of observations for each patient. Given that some patients recorded very few steps on that day (possibly due to limited wear-time), we also compared step counts from the first day and a day with at least 1000 observed steps on the smartphone to allow for a more in-depth assessment of the algorithm,

We created Bland-Altman plots for each dataset and all of the datasets were combined within each validation scheme. Mean bias and limits of agreement (LoA) were calculated to describe the level of agreement between step counts. Mean bias was calculated as the mean difference between two methods of measurement, while LoA were calculated as the mean difference ± 1.96 standard deviation (SD). Participant demographics, body measures, and step count statistics were reported as a range and mean ± SD, whenever available.

Step counts were calculated in Python using previously published open-source method (31). Statistical analysis and visualization were prepared in MATLAB (R2022a; MathWorks, Natick, MA).

## Results

### Internal validation

The aggregated internal validation dataset consisted of data from healthy 103 participants (66 males, representing 64% of the dataset) between 16 and 62 years of age (25.2±7.1). All datasets, except for *PARUSS* provided data on participants’ height and weight, which ranged between 151 and 196 cm (173.8±8.5), and 47 and 112 kg (72.2±14.7), respectively. Participants’ body mass index (BMI) ranged between 17 and 35 kg/m^2^ (23.8±4.1).

In this validation, step counts were aggregated separately for each walking bout across different body locations, including the thigh (n=83 bouts), waist (n=102), chest (n=51), and arm (n=25). Cumulatively, we examined 232 sensor body location pairs: thigh vs. waist (n=83), thigh vs. chest (n=32), thigh vs. arm (n=25), waist vs. chest (n=51), waist vs. arm (n=25), and chest vs. arm (n=15).

On average, in the aggregated internal validation dataset participants performed a mean of 751.7±581.2 steps per walking bout. Mean step counts varied by the dataset; e.g., participants’ mean step counts were 501.5±127.2 in *DaLiAc*, 337.5±14.6 steps in *PARUSS*, 1007.2±79.6 steps in *RealWorld*, 14.6±1.7 steps in *SFDLA*, and 1408.7±561.5 steps in *SPADES*.

**Figure 2a** displays the Bland-Altman plots for the aggregated internal validation dataset. Comparisons between individual studies are provided in **Supplementary Figure 1a-e**. Across the aggregated dataset, the mean bias was equal to -7.2 steps (LoA -47.6, 33.3) or -0.5 %. The largest relative overestimation observed was between the waist and chest in *SFDLA*, and equaled 1.2 steps (LoA -4.3, 6.8) or 8.5 % of the total steps. The largest underestimation was observed between the thigh and waist in *SPADES*, and equaled -28.7 steps (LoA -107.1, 49.7) or -2.0 % of the total steps.

**Figure 2.**
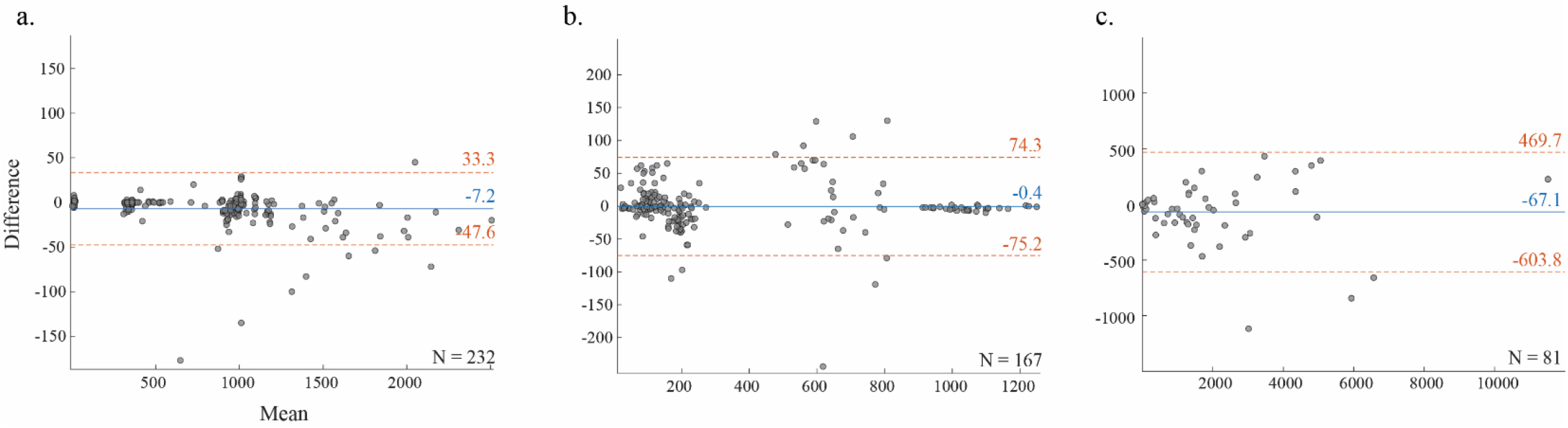
Bland-Altman plots with comparison of step counts in three validation approaches: internal (a.), manual (b.), and wearable (c.). The horizontal axis indicates a mean step count from the two body locations (a.), estimated steps and manually counted ground truth (b.), and estimated steps and step counts obtained from Fitbit (c.). The vertical axis indicates a difference between step counts from the two methods. Blue solid lines indicate mean bias while dashed red lines indicate ±95% limits of agreement calculated as ±1.96 of standard deviations of the differences between the two methods.

### Manual validation

The manual validation of our method included 107 healthy participants. Demographic and anthropometric measurements were only available in *PedEval*. This dataset combined 30 participants, 15 of whom were males, whose ages ranged between 19 and 27 years of age (21.9±52.4) whose heights ranged between 152 and 193 cm (171.0±10.8), and weights ranged between 43 and 136 kg (70.5±17.6). Participants’ BMIs ranged between 17 and 37 kg/m^2^ (23.8±3.7).

We estimated the step count bias based upon 167 comparisons, including 77 comparisons from the *WalkRec* dataset and 90 from the *PedEval* dataset (30 per task). Participants’ mean step count in the aggregated manual validation dataset was 367.4±359.4 steps according to the ground truth (**Figure 2b**). *WalkRec* participants’ means steps were 126.8±59.2 steps, while *PedEval* participants’ steps varied by activity and were 1025.0±171.3 steps in task 1, 648.5±126.3 steps in task 2, and 179.2±22.7 steps in task 3 (**Supplementary Figure 1f-g**). The corresponding estimations calculated using our method were a mean of 119.8±62.2 steps for *WalkRec*, and 1027.5±175.0 steps for task 1, 641.1±137.3 steps for task 2, and 210.8±18.7 steps for task 3. The mean bias across the aggregated dataset, was -0.4 steps (LoA -75.2, 74.3) or 0.1 %. The largest relative overestimation was +8.8 steps (LoA -32.1, 49.7) or 6.9 % within the *WalkRec* dataset. The largest underestimation was -32.3 steps (LoA, - 80.4, 15.8) or -18 %, observed in task 3 in the *PedEval* dataset.

### Wearable validation

Our wearable validation included data from 45 female patients advanced gynecological cancers. Their ages ranged between 24 and 79 years of age (61.5±11.8). Their heights ranged between 148 and 172 cm (159.9±6.1), weights ranged between 48 and 107 kg (67.8±13.0), and BMIs ranged between 19 and 43 kg/m^2^ (23.8±3.7).

Our Bland-Altman analysis included over 81 observations of daily step counts (**Figure 1c**), involving 45 days that constituted the first full day of observation (**Supplementary Figure 1g**) and 36 first days with at least 1000 steps estimated from a smartphone (**Supplementary Figure 1h**). Nine participants did not have any days with more than 1000 steps observed, likely due to limited smartphone wear-time. In the aggregated dataset, the algorithm estimated a mean daily step count of 1998.2±2350.3 steps, which included a mean daily step count of 1371.3±2343.1 steps observed during the first day and 2816.7±2123.6 steps during the first day with at least 1000 steps observed. Comparisons with data from the Fitbit were similar, including a mean daily step count of 1931.2±2338.4 across participants, a mean daily step count of 1316.4±2320.2 steps during the first day, and a mean daily step count of 2733.7±2136.9 steps during the first day with at least 1000 steps observed, respectively. The aggregated estimation bias of the smartphone versus the Fitbit was -67.1 steps (LoA -603.8, 469.7) or 0.3 %, with an underestimation of -54.9 steps (LoA -485.3, 375.6) or -0.2 % during the first day, and -83.0 steps (LoA - 738.5, 572.6) or -0.5 % during the first day with at least 1000 steps.

## Discussion

The limitations of wearable devices, particularly commercial activity trackers, have motivated public health researchers to seek alternative means to quantify human activities. While commercial wearable activity trackers are limited by proprietary summaries of the data, often provided at a daily or hourly level, smartphones offer an opportunity to access raw sub-second level data which can be used to generate reproducible and interpretable digital biomarkers, such as step counts, with research software. Moreover, smartphones are typically carried close to the center of the body, which produces less noisy inertial sensor data compared with the locations that many available wearables are worn (i.e., on the wrist), which provides more accurate estimations of whole-body motion (18,19,41–44).

In this paper, we validated the performance of step counting using a previously published open-source walking recognition method for body-worn devices that contain an accelerometer (29). This method leverages the observation that regardless of sensor location on the body, during walking activity the predominant component of the accelerometer signal transformed to the frequency domain, i.e., step frequency, remains the same, enabling the calculation of the number of steps a person performed in a given time fragment. In our previous study, we validated this approach for walking recognition using data from 1240 subjects gathered in 20 publicly available datasets, and demonstrated that our method estimates walking periods with high sensitivity and specificity: the average sensitivity ranged between 0.92 and 0.97 across various body locations, and the average specificity was largely above 0.95 for common daily activities (household activities, using motorized transportation, cycling, running, desk work, sedentary periods, eating, drinking). Importantly, the method’s performance was not sensitive to different demographics and metrological factors for individual subjects or studies, including participants’ ages, sexes, heights, weights, BMIs, sensor body locations, and measurement environments.

In this study, we further extend this work by validating the performance of step counting method using data collected from 255 subjects in eight independent studies with three goals in mind: (1) to assess the concordance of step counts across various body locations, (2) to compare the method’s estimates with observed step counts, and (3) to compare the method’s estimates with indications of commercial activity tracker (Fitbit Charge 2). The first comparison, an internal validation, demonstrated very high agreement between step counts measured from smartphones locate at most of the places where smartphones are typical worn, i.e., thigh, waist, chest, and arm. This result suggests that our method can be used to assess steps without restricting where participants wear their smartphones, which may reduce participant burden during data collection and help improve long-term study adherence.

Our manual validation on uninterrupted walking revealed almost perfect agreement between the step counts estimated with our method and those denoted by a visual observer. In this case, the absolute difference observed between the two measures was consistently below 1 % (**Supplementary Figure 1g**), which is similar to the results achieved with deep learning methods validated on this dataset in the past (45,46). These results reinforce the utility of using this method in controlled conditions, e.g., to evaluate participants’ functional capacity using a 6-minute walk test, and indicates that the method provides highly accurate estimation of step counts across various sensor locations during regular flat walking.

The mean step counting bias was also low for semi-controlled walking tasks recorded in *PedEval*, free-living tasks recorded in *WalkRec*, and for both scenarios within the wearable validation (first day, first day with at least 1000 steps). In these instances, however, the analysis revealed wider limits of agreement, which may result from a more complex structure of the underlying data which involved walking only few steps at a time as well as sudden changes in walking direction and altitude (e.g., stair climbing) (47). As discussed previously (29), in walking signals with such characteristics the step frequency tends to be modulated by it sub- and higher harmonics which might be identified as dominant in the wavelet decomposition outcome, and mislead our method.

Even more challenging data were analyzed in the wearable validation cohort. Here, the data were collected at unspecified locations (including novel locations, e.g., a bag or backpack), and included data representing various activities of daily living, such as grocery shopping, riding in a car, and doing dishes which might artificially inflate the estimated step counts by either method. This is a likely reason why the comparisons had wider discrepancies, even after removing minutes with 0 steps recorded by either device. Nevertheless, the estimated bias remained low, which indicates that our validated method provides reliable step count estimates across populations and conditions.

Our analysis has several limitations that should be addressed in future studies. First, due to the lack of available datasets, our method was not validated in individuals with walking impairments or those requiring walking aids, such as cane or walkers. Similarly, this method has not been validated in children and many elders, although the mean age of participants in our wearable validation set was 61.5 years and 10% were above 74 years of age.

Further research is needed to understand the frequency-domain gait characteristics in the presence of limping, as well as the potential overlap between the step frequency of walking activity in children and that of running activity in adults (48,49). The latter might be particularly important in studies that differentiate steps performed during leisure and exertional activities. Second, the wearable validation was performed with the use of a proprietary activity tracker (Fitbit Charge 2). Although this device has demonstrated reliable step counts during naturalistic gait performed in laboratory conditions (50,51), its accuracy in free-living conditions is inconclusive, and it is presumably dependent on the characteristics of the studied population (20,52,53). Importantly, the selected activity tracker was placed on the wrist, a body location that can be activated by many repetitive movements (e.g., gesticulating) while the rest of the body is still, hence it is more likely to overestimate steps compared to locations closer to the body mass center. To improve comparisons with our method, in wearable validation we removed data instances when either method indicated 0 steps. Finally, the estimation of step counts in free-living studies must account for non-wear time of smartphones (e.g., while the phone is charging or sitting on a table). Unlike many wearables that are attached to the body (e.g., wristbands), smartphones can be easily set aside, sometimes for prolonged periods of time. Such situations introduce considerable discrepancy between the estimated and actual number of steps a person performs during the day and should be reported, ideally with confidence intervals.

In conclusion, we performed a three-way validation of a robust, reproducible, and scalable method for step counting using smartphones and other wearable activity trackers. This validation demonstrates that our approach provides reliable step counts across sensor locations and populations, including healthy adults and those with incurable cancers. The method performed well in multiple environments, including indoors, outdoors, and in day-to-day life across settings. This method is a promising strategy for studying human gait with personal smartphones which does not require active patient participation or the introduction of new devices.

## Data Availability

All data produced in the present study are available upon reasonable request to the authors.

## Acknowledgements

We would like to thank the patients in the *HOPE* study for their dedication in data collection. We would also like to thank the researchers for making their datasets publicly available.

Drs Straczkiewicz and Onnela are supported by NHLBI award U01HL145386 and NIMH award R37MH119194. Drs Wright and Onnela are also supported by NINR award R21NR018532. Dr. Matulonis is supported by the Dana-Farber Harvard Cancer Center Ovarian Cancer SPORE grant (P50CA240243) and the Breast Cancer Research Foundation.

MS – Study concept and design, method development, data collection, data processing, data analysis, figure and table preparation, manuscript drafting.

NLK – Study concept and design, data collection, critical review of manuscript.

ET – Data collection, critical review of manuscript.

UM – Data collection, critical review of manuscript.

SMC – Data collection, critical review of manuscript.

AAW – Study concept and design, data collection, data analysis, critical review of manuscript, scientific supervision.

JPO – Study concept and design, method development, data collection, data analysis, critical review of manuscript, scientific supervision.

## Abbreviations

BMI: Body mass index
DaLiAc: Daily life activities dataset
HOPE: Helping our patients excel dataset
LoA: Limits of agreement
PARUSS: Physical activity recognition using smartphone sensors dataset
PedEval: Pedometer evaluation project dataset
RealWorld: Real-world dataset
SFDLA: Simulated falls and daily living activities dataset
SPADES: Human physical activity dataset
WalkRec: Walking recognition dataset

## References

1. U.S. Department of Health and Human Services. Step It Up! The Surgeon General’s Call to Action to Promote Walking and Walkable Communities [Internet]. Washington, DC; 2015. Available from: www.surgeongeneral.gov

2. Hanson S, Jones A. Is there evidence that walking groups have health benefits? A systematic review and meta-analysis. Br J Sports Med [Internet]. 2015 Jun 1;49(11):710 LP –715. Available from: http://bjsm.bmj.com/content/49/11/710.abstract

3. Harris T, Limb ES, Hosking F, Carey I, DeWilde S, Furness C, et al. Effect of pedometer-based walking interventions on long-term health outcomes: Prospective 4-year follow-up of two randomised controlled trials using routine primary care data. PLoS Med. 2019 Jun;16(6):e1002836.

4. Paluch AE, Bajpai S, Bassett DR, Carnethon MR, Ekelund U, Evenson KR, et al. Daily steps and all-cause mortality: a meta-analysis of 15 international cohorts. Lancet Public Heal. 2022 Mar;7(3):e219–28.

5. Jefferis BJ, Whincup PH, Papacosta O, Wannamethee SG. Protective effect of time spent walking on risk of stroke in older men. Stroke. 2014 Jan;45(1):194–9.

6. Hall KS, Hyde ET, Bassett DR, Carlson SA, Carnethon MR, Ekelund U, et al. Systematic review of the prospective association of daily step counts with risk of mortality, cardiovascular disease, and dysglycemia. Int J Behav Nutr Phys Act [Internet]. 2020;17(1):78. Available from: https://doi.org/10.1186/s12966-020-00978-9

7. Kraus WE, Janz KF, Powell KE, Campbell WW, Jakicic JM, Troiano RP, et al. Daily Step Counts for Measuring Physical Activity Exposure and Its Relation to Health. Med Sci Sports Exerc. 2019 Jun;51(6):1206–12.

8. Master H, Annis J, Huang S, Beckman JA, Ratsimbazafy F, Marginean K, et al. Association of step counts over time with the risk of chronic disease in the All of Us Research Program. Nat Med [Internet]. 2022;28(11):2301–8. Available from: https://doi.org/10.1038/s41591-022-02012-w

9. del Pozo Cruz B, Ahmadi MN, Lee I-M, Stamatakis E. Prospective Associations of Daily Step Counts and Intensity With Cancer and Cardiovascular Disease Incidence and Mortality and All-Cause Mortality. JAMA Intern Med [Internet]. 2022 Nov 1;182(11):1139–48. Available from: https://doi.org/10.1001/jamainternmed.2022.4000

10. Bennett A V, Reeve BB, Basch EM, Mitchell SA, Meeneghan M, Battaglini CL, et al. Evaluation of pedometry as a patient-centered outcome in patients undergoing hematopoietic cell transplant (HCT): a comparison of pedometry and patient reports of symptoms, health, and quality of life. Qual life Res an Int J Qual life Asp Treat care Rehabil. 2016 Mar;25(3):535–46.

11. Coorevits L, Coenen T. The rise and fall of wearable fitness trackers. Acad Manag. 2016;

12. Wigginton C. Global mobile consumer trends, 2nd edition [Internet]. 2017. Available from: https://www2.deloitte.com/global/en/pages/technology-media-and-telecommunications/articles/gx-global-mobile-consumer-trends.html

13. Mercer K, Giangregorio L, Schneider E, Chilana P, Li M, Grindrod K. Acceptance of commercially available wearable activity trackers among adults aged over 50 and with chronic illness: a mixed-methods evaluation. JMIR mHealth uHealth. 2016;4(1):e7.

14. Hermsen S, Moons J, Kerkhof P, Wiekens C, De Groot M. Determinants for Sustained Use of an Activity Tracker: Observational Study. JMIR mHealth uHealth. 2017 Oct;5(10):e164.

15. Rubin DS, Ranjeva SL, Urbanek JK, Karas M, Madariaga MLL, Huisingh-Scheetz M. Smartphone-Based Gait Cadence to Identify Older Adults with Decreased Functional Capacity. Digit Biomarkers [Internet]. 2022;6(2):61–70. Available from: https://www.karger.com/DOI/10.1159/000525344

16. Major MJ, Alford M. Validity of the iPhone M7 motion co-processor as a pedometer for able-bodied ambulation. J Sports Sci. 2016 Dec;34(23):2160–4.

17. Karas M, Bai J, Strączkiewicz M, Harezlak J, Glynn NW, Harris T, et al. Accelerometry Data in Health Research: Challenges and Opportunities: Review and Examples. Stat Biosci. 2019;11(2):210–237.

18. Atallah L, Lo B, King R, Yang G-Z. Sensor Positioning for Activity Recognition Using Wearable Accelerometers. IEEE Trans Biomed Circuits Syst. 2011;5(4):320–9.

19. Kooiman TJM, Dontje ML, Sprenger SR, Krijnen WP, van der Schans CP, de Groot M. Reliability and validity of ten consumer activity trackers. BMC Sport Sci Med Rehabil. 2015;7:24.

20. Collins JE, Yang HY, Trentadue TP, Gong Y, Losina E. Validation of the Fitbit Charge 2 compared to the ActiGraph GT3X+ in older adults with knee osteoarthritis in free-living conditions. PLoS One. 2019;14(1):e0211231.

21. Onnela J-P, Rauch SL. Harnessing Smartphone-Based Digital Phenotyping to Enhance Behavioral and Mental Health. Neuropsychopharmacology [Internet]. 2016;41(7):1691–6. Available from: https://doi.org/10.1038/npp.2016.7

22. Harari GM, Müller SR, Aung MSH, Rentfrow PJ. Smartphone sensing methods for studying behavior in everyday life. Curr Opin Behav Sci [Internet]. 2017;18:83–90. Available from: https://www.sciencedirect.com/science/article/pii/S2352154617300505

23. Mohr DC, Zhang M, Schueller SM. Personal Sensing: Understanding Mental Health Using Ubiquitous Sensors and Machine Learning. Annu Rev Clin Psychol. 2017 May;13:23–47.

24. Straczkiewicz M, James P, Onnela J-P. A systematic review of smartphone-based human activity recognition methods for health research. npj Digit Med [Internet]. 2021;4(1):148. Available from: https://doi.org/10.1038/s41746-021-00514-4

25. Onnela J-P. Opportunities and challenges in the collection and analysis of digital phenotyping data. Neuropsychopharmacology [Internet]. 2021;46(1):45–54. Available from: https://doi.org/10.1038/s41386-020-0771-3

26. Anderson M, Perrin A. Tech Adoption Climbs Among Older Adults [Internet]. Pew Research Center; 2017. Available from: http://www.pewinternet.org/wp-content/uploads/sites/9/2017/05/PI_2017.05.17_Older-Americans-Tech_FINAL.pdf

27. Yang R, Wang B. PACP: A position-independent activity recognition method using smartphone sensors. Inf [Internet]. 2016;7(4):72. Available from: https://www.scopus.com/inward/record.uri?eid=2-s2.0-85007346895&doi=10.3390%2Finfo7040072&partnerID=40&md5=11e596de656775b5cf403967261cb2e 6

28. Saha J, Chowdhury C, Chowdhury IR, Biswas S, Aslam N. An ensemble of condition based classifiers for device independent detailed human activity recognition using smartphones. Inf [Internet]. 2018;9(4):94. Available from: https://www.scopus.com/inward/record.uri?eid=2-s2.0-85045750633&doi=10.3390%2Finfo9040094&partnerID=40&md5=3daae1109db607a71cfa15193e14089 c

29. Straczkiewicz M, Huang EJ, Onnela J-P. A “one-size-fits-most” walking recognition method for smartphones, smartwatches, and wearable accelerometers. npj Digit Med [Internet]. 2023;6(1):29. Available from: https://doi.org/10.1038/s41746-022-00745-z

30. The method proposed in this paper is available as a find_walking function: https://github.com/MStraczkiewicz/find_walking (MATLAB).

31. The method used in this paper is available as the Oak tree within the Forest library: https://github.com/onnela-lab/forest (Python).

32. Leutheuser H, Schuldhaus D, Eskofier BM. Hierarchical, Multi-Sensor Based Classification of Daily Life Activities: Comparison with State-of-the-Art Algorithms Using a Benchmark Dataset. PLoS One [Internet]. 2013;8(10):1–11. Available from: https://doi.org/10.1371/journal.pone.0075196

33. Shoaib M, Bosch S, Durmaz Incel O, Scholten H, Havinga PJM. Fusion of smartphone motion sensors for physical activity recognition. Sensors (Switzerland) [Internet]. 2014;14(6):10146–76. Available from: https://www.scopus.com/inward/record.uri?eid=2-s2.0-84902254076&doi=10.3390%2Fs140610146&partnerID=40&md5=a0e6cdf165a47c4c13937d6f861bee60

34. Sztyler T, Stuckenschmidt H. On-body localization of wearable devices: An investigation of position-aware activity recognition. In: 2016 IEEE International Conference on Pervasive Computing and Communications (PerCom). 2016. p. 1–9.

35. Özdemir AT, Barshan B. Detecting Falls with Wearable Sensors Using Machine Learning Techniques. Sensors [Internet]. 2014;14(6):10691–708. Available from: https://www.mdpi.com/1424-8220/14/6/10691

36. John D, Tang Q, Albinali F, Intille S. An Open-Source Monitor-Independent Movement Summary for Accelerometer Data Processing. J Meas Phys Behav [Internet]. 2(4):268–81. Available from: http://journals.humankinetics.com/view/journals/jmpb/2/4/article-p268.xml

37. Casado FE, Rodríguez G, Iglesias R, Regueiro C V, Barro S, Canedo-Rodríguez A. Walking Recognition in Mobile Devices. Vol. 20, Sensors. 2020.

38. Mattfeld R, Jesch E, Hoover A. A new dataset for evaluating pedometer performance. In: 2017 IEEE International Conference on Bioinformatics and Biomedicine (BIBM). 2017. p. 865–9.

39. Wright AA, Raman N, Staples P, Schonholz S, Cronin A, Carlson K, et al. The HOPE Pilot Study: Harnessing Patient-Reported Outcomes and Biometric Data to Enhance Cancer Care. JCO Clin cancer informatics. 2018 Dec;2:1–12.

40. Onnela J-P, Dixon C, Griffin K, Jaenicke T, Minowada L, Esterkin S, et al. Beiwe: A data collection platform for high-throughput digital phenotyping. J Open Source Softw. 2021;6(68):3417.

41. Straczkiewicz M, Glynn NW, Zipunnikov V, Harezlak J. Fast and Robust Algorithm for Detecting Body Posture Using Wrist-Worn Accelerometers. J Meas Phys Behav [Internet]. 2020;3(4):285–93. Available from: https://journals.humankinetics.com/view/journals/jmpb/3/4/article-p285.xml

42. Tudor-Locke C, Barreira T V, Schuna JMJ. Comparison of step outputs for waist and wrist accelerometer attachment sites. Med Sci Sports Exerc. 2015 Apr;47(4):839–42.

43. Mandigout S, Lacroix J, Perrochon A, Svoboda Z, Aubourg T, Vuillerme N. Comparison of Step Count Assessed Using Wrist- and Hip-Worn Actigraph GT3X in Free-Living Conditions in Young and Older Adults. Front Med [Internet]. 2019;6. Available from: https://www.frontiersin.org/articles/10.3389/fmed.2019.00252

44. Nelson RK, Hasanaj K, Connolly G, Millen L, Muench J, Bidolli NSC, et al. Comparison of Wrist- and Hip-Worn Activity Monitors When Meeting Step Guidelines. Prev Chronic Dis. 2022;19:210343.

45. Luu L, Pillai A, Lea H, Buendia R, Khan FM, Dennis G. Accurate Step Count with Generalized and Personalized Deep Learning on Accelerometer Data. Sensors (Basel). 2022 May;22(11).

46. Khan SS, Abedi A. Step counting with attention-based LSTM.

47. Mattfeld R, Jesch E, Hoover A. Evaluating Pedometer Algorithms on Semi-Regular and Unstructured Gaits. Sensors (Basel). 2021 Jun;21(13).

48. Davis JJ, Straczkiewicz M, Harezlak J, Gruber AH. CARL: a running recognition algorithm for free-living accelerometer data. Physiol Meas [Internet]. 2021;42(11):115001. Available from: http://dx.doi.org/10.1088/1361-6579/ac41b8

49. Tudor-Locke C, Schuna JM, Han H, Aguiar EJ, Larrivee S, Hsia DS, et al. Cadence (steps/min) and intensity during ambulation in 6–20 year olds: the CADENCE-kids study. Int J Behav Nutr Phys Act [Internet]. 2018;15(1):20. Available from: https://doi.org/10.1186/s12966-018-0651-y

50. Curran M, Tierney A, Collins L, Kennedy L, McDonnell C, Sheikhi A, et al. Accuracy of the ActivPAL and Fitbit Charge 2 in measuring step count in Cystic Fibrosis. Physiother Theory Pract. 2022 Nov;38(13):2962–72.

51. Roberts-Lewis SF, White CM, Ashworth M, Rose MR. Validity of Fitbit activity monitoring for adults with progressive muscle diseases. Disabil Rehabil [Internet]. 2022;44(24):7543–53. Available from: https://doi.org/10.1080/09638288.2021.1995057

52. Tedesco S, Sica M, Ancillao A, Timmons S, Barton J, O’Flynn B. Validity Evaluation of the Fitbit Charge2 and the Garmin vivosmart HR+ in Free-Living Environments in an Older Adult Cohort. JMIR Mhealth Uhealth [Internet]. 2019 Jun;7(6):e13084. Available from: http://www.ncbi.nlm.nih.gov/pubmed/31219048

53. Irwin C, Gary R. Systematic Review of Fitbit Charge 2 Validation Studies for Exercise Tracking. Transl J Am Coll Sport Med [Internet]. 2022;7(4). Available from: https://journals.lww.com/acsm-tj/Fulltext/2022/10140/Systematic_Review_of_Fitbit_Charge_2_Validation.8.aspx

