## Supplementary Figure 1 for "Validation of an open-source smartphone step counting algorithm in clinical and non-clinical settings"

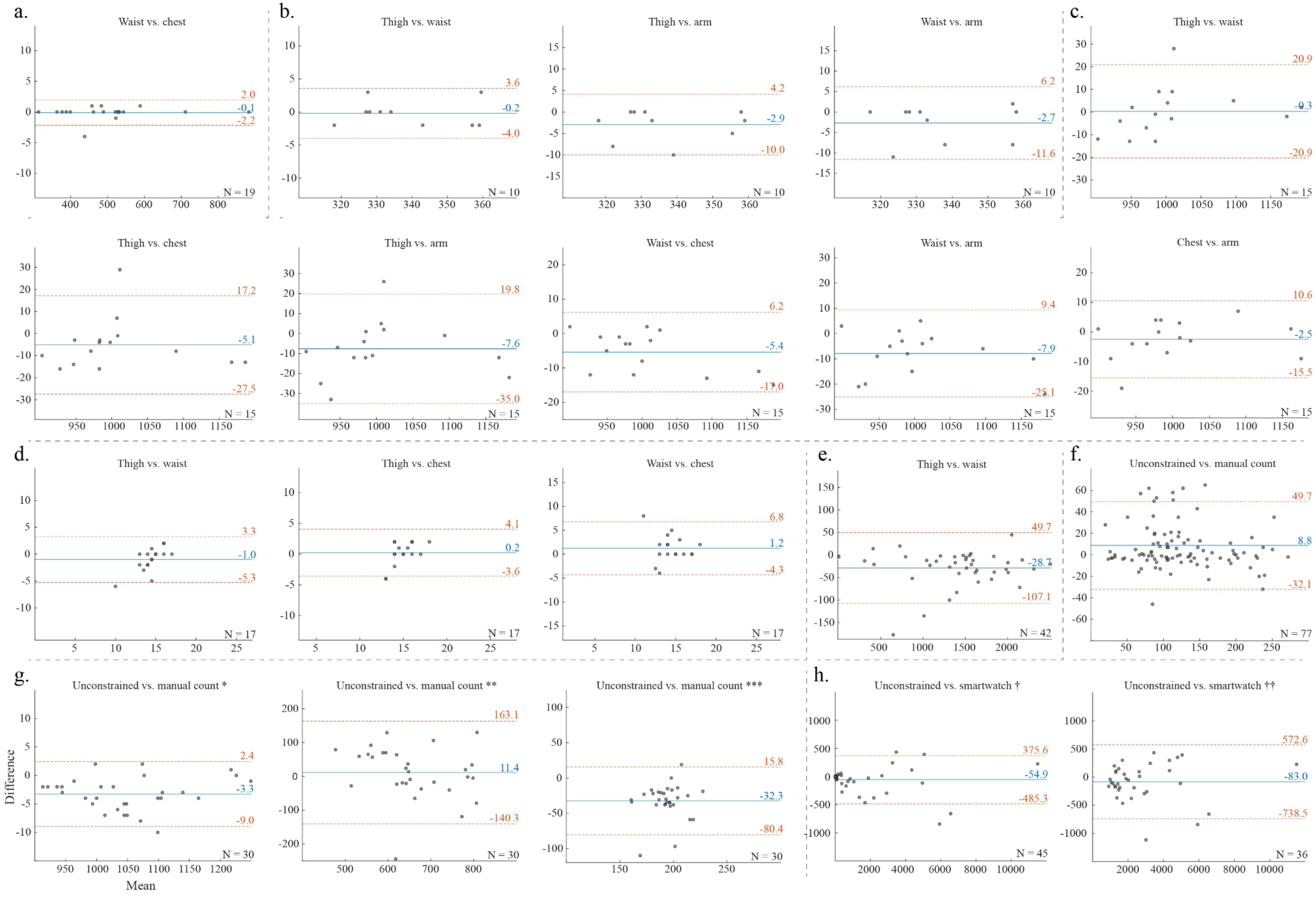


**Supplementary Figure 1.** Bland-Altman plots with comparison between step estimates in the included studies: *DaLiAc* (a.), *PARUSS* (b.), *RealWorld* (c.), *SFDLA* (d.), *SPADES* (e.), *WalkRec* (f.), *PedEval* (g.), and *HOPE* (h.). In *PedEval*, * indicates task 1, ** indicates task 2, and *** indicates task 3. In *HOPE*, † indicates analysis over the first full day past enrollment and †† indicates analysis over the first full day with more than 1000 steps. Algorithm performance was compared using step estimates from various body locations (a.-e.), manual annotation (f.-g.), and Fitbit (h.).
